# The Safety, Effectiveness and Prognostic Factors of Intravitreal Avastin (Bevacizumab) Injection for the Treatment of Macular Edema at the University of Gondar Tertiary Eye Care and Training Center, NW Ethiopia

**DOI:** 10.1101/2024.07.10.24310159

**Authors:** Yoseph Gizachew, Asamere Tsegaw, Tarekegn Wuletaw

## Abstract

**Introduction:** The current management of macular edema (ME) is intravitreal injection of anti-vascular endothelial growth factor (anti-VEGF) drugs and this represents an important advance in the treatment of ME.

Studies done in the western eye care settings have confirmed that intravitreal injection of Avastin is effective for the treatment of ME. However, data on this drug’s efficacy and safety in African eye care settings are very scarce.

**Objective:** To assess the Safety and Effectiveness of intravitreal Bevacizumab (Avastin) injection for the treatment of Macular Edema (ME) due to retina vascular diseases at university of Gondar tertiary eye care and training center, NW Ethiopia.

**Method:** A retrospective study was done on patients who were given intravitreal avastin (IVA) for the treatment of diabetic macular edema (DME), retinal vein occlusion (RVO) and Neovascular Age related macular degeneration (AMD). The main outcome measure was visual acuity (VA) and central macular thickness (CMT) measured by spectral domain OCT.

**Results:** Medical records of 50 patients (66 eyes) were reviewed of which 46 (69.7%) were males and mean age of 54.2 years (range 20-80). The means of baseline VA and CMT were 1.0logMAR and 379.4 µm respectively. At the end of follow up and after mean injection of 2.5 times per eye, the mean VA improved to 0.7 logMAR (p=0.001) and the mean CMT reduced to 295 µm (p=0.0001). Baseline mean VA was significant prognostic factor for VA improvement (p=0.0001). Baseline mean CMT (P=0.007), number of injection (P=0.009) and diffuse macular edema (P=0.03) were significant factors for CMT reduction.

**Conclusions:** IVA injection for ME edema due to retinal vascular diseases resulted in a significant improvement in mean VA (p=0.001) and CMT (p=0.0001) at the end of follow up. There was no any ocular or systemic complication of IVA injection.

## Introduction

Macular edema (ME) is an abnormal thickening of the macula due to the accumulation of excess fluid in the extracellular space of the retina. ME occurs as a result of abnormal retinal vascular permeability and break down of the blood-retinal barrier mainly mediated by vascular endothelial growth factors (VEGF) and other cytokines (1).

ME is caused by a variety of ophthalmic conditions such as diabetic retinopathy (DR), retinal vein occlusion (RVO) and intraocular inflammation (uveitis) (1).

Among patients with DR, diabetic macular edema (DME) is the most frequent cause of vision impairment(2). Macular edema (ME) is also the main cause of decreased visual acuity (VA) after retinal vein occlusion and neovascular AMD (3).

Currently VEGF inhibitors are the first-line of treatment for macular edema and intraocular neovascularization(1). Their excellent safety profile and lower side effects make anti VEGF agents the current recommended first-line therapy for ME and intraocular neovascularization (4).

Avastin is a recombinant humanized monoclonal IgG1 antibody that binds to and inhibits the biologic activity of human VEGF and currently used for intravitreal administration for the treatment of wet AMD, DME and ME due to RVO (1).

Several studies done mainly in western eye care settings have confirmed that intravitreal injection of Avastin was safe and effective (5–16).

However, Avastin is still being used as an off-label drug and evidences on its effectiveness and safety in African eye care settings are very scarce (17).

This is the first study done in Ethiopia to assess the safety, effectiveness (both anatomical and functional outcome) and prognostic factors of intravitreal Bevacizumab (Avastin) injection for the treatment of Macular Edema.

## Materials and Methods

### Study design and period

A hospital based retrospective cross-sectional study was done on patients who were given intravitreal Avastin at the University of Gondar Tertiary eye care and training center from March 1st 2022 to November 30th 2022.

### Study area

The study was conducted at University of Gondar Tertiary Eye Care and Training Center, a major eye care and training center in Ethiopia. It is an ophthalmic referral center for an estimated 20 million people living in North-West Ethiopia. The center provides eye care services both at base hospital and rural outreach sites. The base hospital has 8 out-patient clinics, facilities for in-patient care with 30 beds and five operation theatres.

Currently there are 5 subspecialty clinics with 8 actively working ophthalmologists, 23 ophthalmology trainee residents, 38 optometrists, 35 ophthalmic nurses and general clinical nurses actively working in the outpatient clinics and operation theatres of the tertiary eye care and training center. The retina sub-specialty clinic is active three days a week and has one retina specialist. The retina clinic has patient volume of on average 300 cases per month.

### Sample size and sampling procedure

All consecutive patients who attended UOG comprehensive specialized referral hospital, tertiary eye care and training center retina subspecialty clinic and received intravitreal avastin injection in the study period and fulfill the inclusion criteria was enrolled.

### Inclusion and exclusion criteria

#### Inclusion criteria

All patients who received intravitreal avastin for macular edema due to Diabetic macular edema (DME), Branch RVO (BRVO), Central RVO (CRVO), and wet AMD at UoG comprehensive specialized hospital, tertiary eye care and training center during the study period.

Patients with documented pre-injection macular edema on OCT and CMT ≥ 300micron Baseline Visual acuity ≤ 6/9

#### Exclusion criteria

Patients on follow up after intravitreal Avastin injection elsewhere

Patients who were given intravitreal steroid injection

Patients who were given laser treatment

Patients with no baseline CMT measurement

Patients with ocular comorbidities such as visually significant cataract and corneal opacity, glaucoma, retinal degenerations (myopia, Retinitis pigmentosa etc.), Retinal Detachment, etc

#### Operational definitions

➢ **Improved**--- At least increments of visual acuity (VA) by ≥one line in logMAR (5letters) and decrement of CMT by at least 10% from baseline at 6^th^ month.
➢ **Remain stable** ---- When mean VA is the same or increment by less than 5 letters from baseline and mean CMT reduction less than 10% or increment or decrement by 10% at the end of follow up at the 4th month.
➢ **Worsened**---- Decrement of VA by one-line in logMAR (5 letters) and or CMT is increased by ≥ 10% at 4^th^ month from base-line.

### Data collection tools and procedures

Medical record number of all patients who were given IVA injection was collected from operation room registration book and this number was used to retrieve the medical records of participants from medical record room. Structured questionnaire was used to document relevant information from patient medical records which included socio-demographic data, duration of visual complaint, which eye was involved, diagnosis, and duration of follow up since the first injection, indication for injection and the number of injections was retrieved and recorded. Comorbidities like DM, HTN, and dyslipidemias were documented. Laboratory data such as the most recent fasting blood sugar, lipid profile was documented. Best corrected Visual acuity at the Snellen’s chart, Comprehensive anterior segment examination findings found with slit lamp bio-microscope and detailed examination findings of the posterior segment with 90D volk in a dilated pupil with tropicamide by a senior retina specialist was reviewed. Pre-injection and first day post-injection Intraocular pressure (IOP) in mmHG was measured with iCare® (Tiolat Oy, Helsinki, Finland) tonometer and documented. Central macula thickness (CMT) measured by spectral domain OCT (Meditace, cirrus HD, model 500 CA 84568, USA) at baseline and every month in the follow-up period was recorded in checklist. Number of Avastin injections and complications of injection recorded on medical record were incorporated in the checklist.

A method proposed by Gregori et al. was used to convert VA measured by the Snellen chart into approximate ETDRS letter scores in order to simplify the statistical analysis. The equation used was: Approximate ETDRS letters = 85 + 50 × log (Snellen fraction) (18).

### Data processing and analysis

The collected data was checked for accuracy, completeness and manual data clean up and correction of errors was done. Data was coded and entered into Epi-Data 4.6 and exported to statistical package for social sciences (SPSS) version 25 for further analysis. Results were described in terms of percentages, numbers, means, medians and displayed on tables, bar graphs and pie charts. Paired-samples t-test analysis was used to compare mean VA and mean CMT and P-value of less than 0.05 was considered as statistically significant.

### Ethical considerations

Ethical clearance was obtained from UOG, school of medicine ethical review board (Ref.No. SOM/3092/2023, Date 14/02/2023) and permission to undergo this study was also obtained from the department of ophthalmology. Informed written consent was obtained from patients and it was made clear that the identity of patients and their records was kept confidential and anonymous.

## Results

A total of 66 eyes of 50 participants were included in this study of which 46 (69.7%) were males and 20 (30.3%) were females. The mean age of the study participants was 54.2 ± 16.2years and range between 20 and 80 years. (Table-I)

**Table I:** Baseline clinical characteristics of study eyes that were candidates for IVA injection at UoG tertiary eye care and training center retina sub-specialty clinic, Gondar, Ethiopia, 2023.

| Characters | Number | Percent |
| --- | --- | --- |
| **Gender** |  |  |
| Male | 46 | 69.7 |
| Female | 20 | 30.3 |
| **Comorbidity** |  |  |
| Hypertension | 6 | 9.1 |
| Dyslipidemia | 0 | 0 |
| **Baseline VA** |  |  |
| Mild (6/9 -6/18) | 18 | 27 |
| Moderate (6/18-6/60) | 22 | 33 |
| Severe (6/60-3/60) | 7 | 11 |
| Blind (≤3/60) | 19 | 29 |
| **OCT findings** |  |  |
| CME | 40 | 60 |
| Diffuse ME | 25 | 38 |
| Sub retinal fluid | 1 | 2 |
| **Indication for injection** |  |  |
| NPDR with ME | 22 | 33.3 |
| PDR with ME | 15 | 22.7 |
| BRVO with ME | 7 | 10.6 |
| CRVO with ME | 11 | 16.7 |
| Wet AMD with ME | 11 | 16.7 |

Pre-injection IOP was measured for 51eyes and baseline mean IOP was 15.65±3.4 mmHg (8-25mmHg).

First day post injection IOP was taken for 55 eyes and the mean IOP was 15 ± 3.56 mmHg (8-26mmHg). Only two patients had first day post injection IOP above 20mmHg, one 22mmHg and the second 26mmHg.

The Mean number of intravitreal injections was 2.5 ±0.98 (range 1-5). (Graph-I)

**Graph I:**
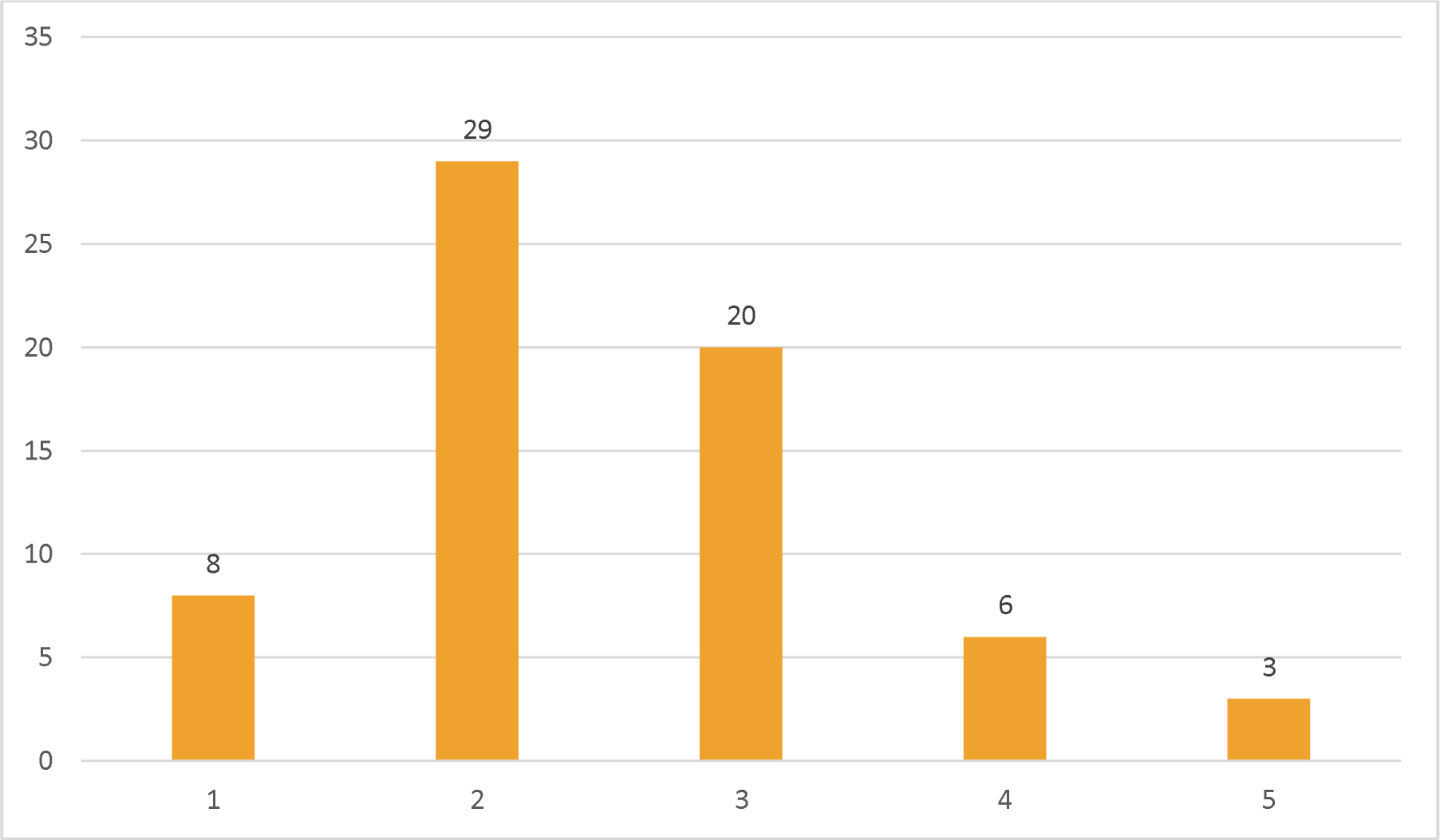
Bar graph showing the number of IVA injections among study eyes at UoG tertiary eye care and training center retina sub-specialty clinic, Gondar, Ethiopia, 2023.

**Graph II:**
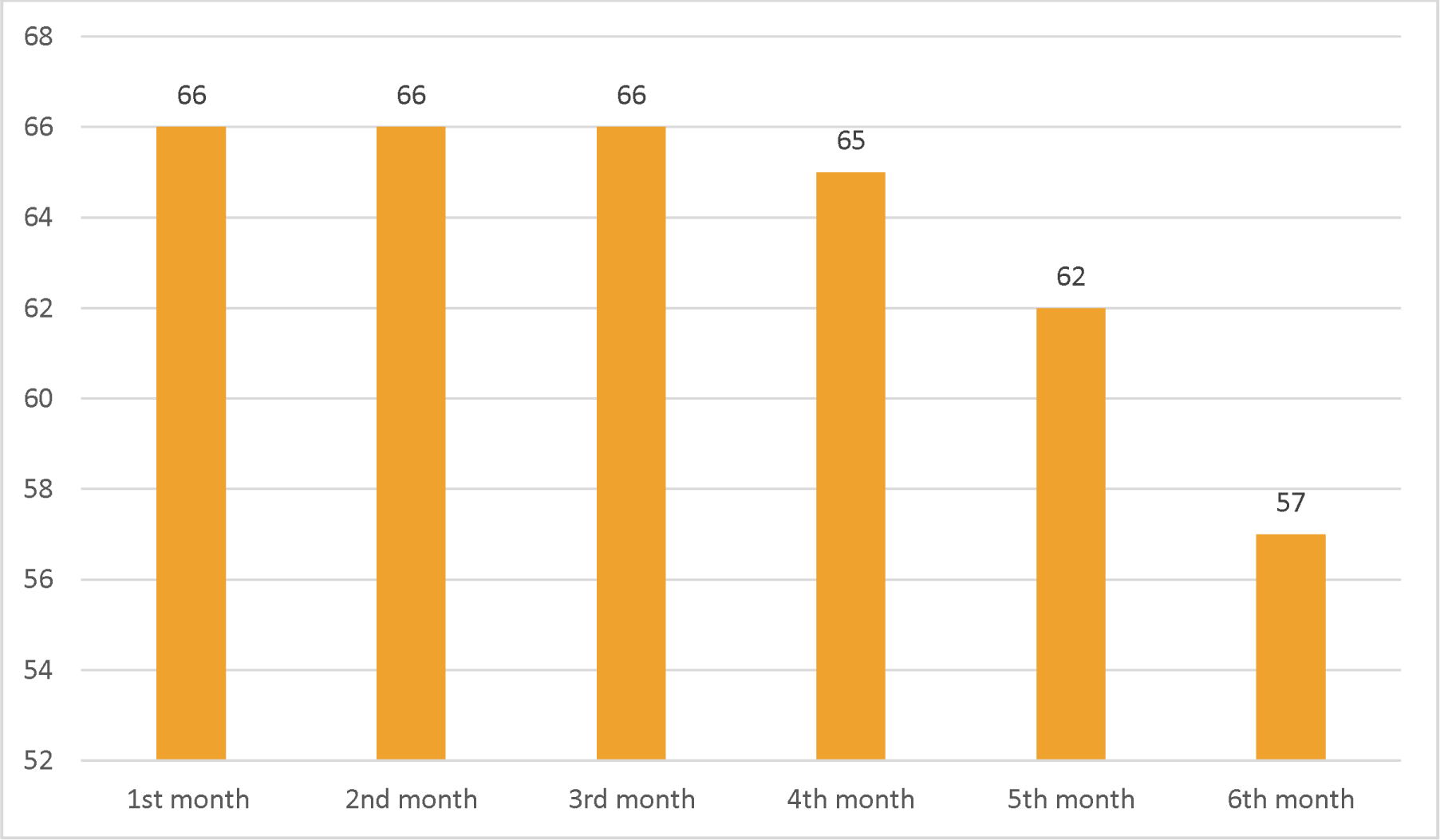
Post IVA injection follow-up visits among study eyes at UoG tertiary eye care and training center retina sub-specialty clinic, Gondar, Ethiopia, 2023

DME was the commonest cause of ME among study patients accounting 56% of which NPDR with DME accounted 33.3% and PDR with DME 22.7%. The second common cause was RVO which accounted 27% of which 16.7% had CRVO and 10.6% had BRVO 10.6%. The rest 16.7% had wet AMD. (Table-I)

At base line, first, second and third month of follow up period CMT was measured for all eyes but at 4^th^ month CMT was measured for 58 eyes. After the 4^th^ month and till the end of follow up only 23 eyes had CMT measurements. (Table-III)

The mean baseline VA was 1±0.7 logMAR (0.18-3) and the mean baseline CMT was 379.44 ± 100.32 µm (300-828µm). (Table II) (Table-III)

**Table II:** Baseline and post injection mean VA in logMAR among study eyes at UoG tertiary eye care and training center retina sub-specialty clinic, Gondar, Ethiopia, 2023

|  |  | Pre-injection VA | 1<sup>ST</sup> month post injection VA | 2<sup>ND</sup> month post injection VA | 3<sup>rd</sup> month post injection VA | 4<sup>th</sup> month post injection VA | 5<sup>th</sup> month post injection VA | 6<sup>th</sup> month post injection VA |
| --- | --- | --- | --- | --- | --- | --- | --- | --- |
| No. | Valid | 66 | 66 | 66 | 66 | 65 | 62 | 57 |
|  | Missing | 0 | 0 | 0 | 0 | 1 | 4 | 9 |
| Mean |  | 1.0552 | .8548 | .8415 | .7832 | .7677 | .7755 | .7053 |
| St.Deviation |  | .73408 | .57283 | .68212 | .57935 | .54156 | .55594 | .49549 |

**Table III:** Baseline and post-injection mean CMT in micrometer of study eyes that were candidates for IVA injection at UoG tertiary eye care and training center retina sub-specialty clinic, Gondar, Ethiopia, 2023

|  |  | Baseline CMT | Post injection 1st month CMT | Post injection 2nd month CMT | Post injection 3rd month CMT | Post injection 4th month CMT | Post injection 5th month CMT | Post injection 6th month CMT |
| --- | --- | --- | --- | --- | --- | --- | --- | --- |
| No. | Valid | 66 | 66 | 66 | 66 | 58 | 20 | 3 |
|  | Missing | 0 | 0 | 0 | 0 | 8 | 46 | 63 |
| Mean CMT |  | 379.44 | 333.47 | 318.82 | 301.83 | 295.17 | 286.50 | 265.33 |
| St.Deviation |  | 100.315 | 67.317 | 67.282 | 62.964 | 66.388 | 55.956 | 63.516 |

After 1 month of IVA injection the mean VA improved from 1.00 logMAR to 0.85 logMAR (p=0.007) which was maintained at second moth. At 3rd month mean VA was increased from 1.00 logMAR to 0.78 (p=0.001) this was maintained throughout the 6 month follow up.(Table-2)

In the sub group analysis patients with DME mean VA improved from 0.79logMAR to 0.63 (P=0.082), to 0.56 (p=0.013), to 0.5 (p=0.002) at 1st month, 3rd month and 6th month respectively. In patients with RVO (CRVO and BRVO) mean VA improved from 1.3logMAR to 0.98(p=0.012), to 0.92(p=0.008), to 0.89(p=0.004) at 1st month, 3rd month and 6th month respectively. In patients with wet AMD mean VA was improved from 1.4logMAR to 1.3(p=0.7), to 1.2 (p=0.33), to 1.0(p=0.33) at 1st month, 3rd month and 6th month respectively. (Graph-III)

**Graph III:**
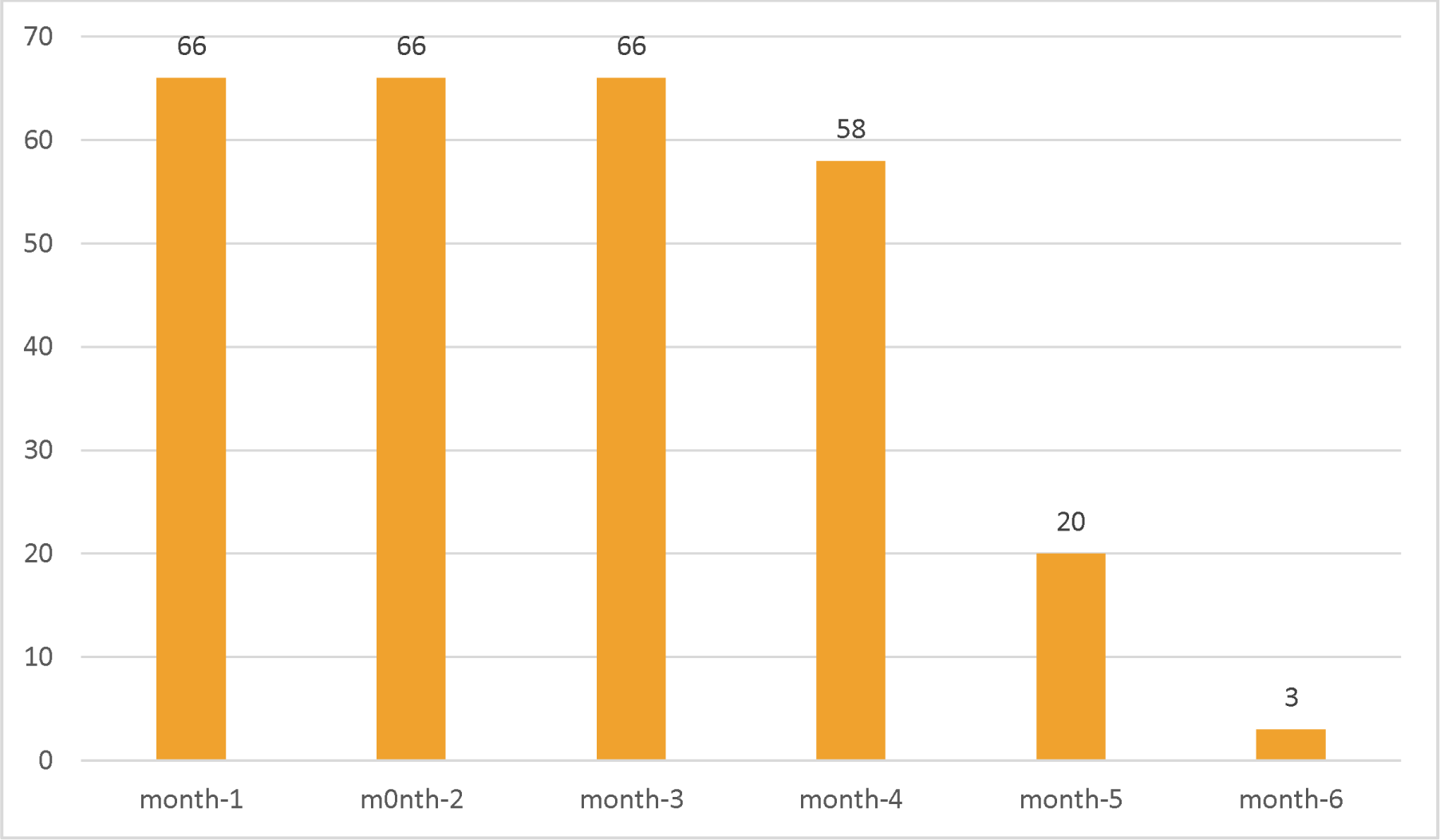
Post IVA injection follow-up visits who had CMT among study eyes at UoG tertiary eye care and training center retina sub-specialty clinic, Gondar, Ethiopia, 2023

**Graph IV:**
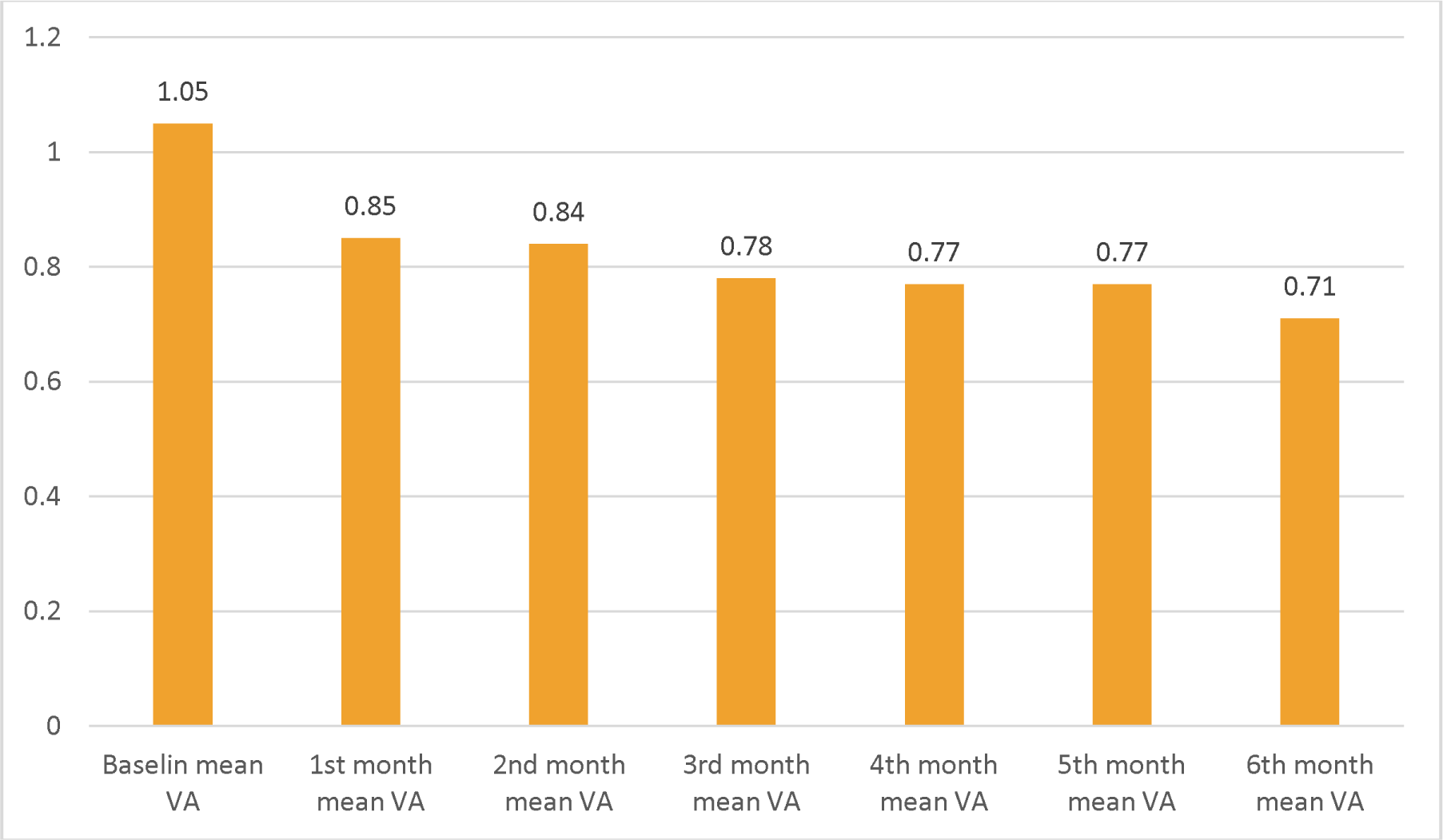
Baseline and post IVA injection mean VA (logMAR) of study eyes that were candidates for IVA injection at UoG tertiary eye care and training center retina sub-specialty clinic, Gondar, Ethiopia, 2023.

After IVA injection the mean CMT decreased from baseline 379.44 µm to 333.47µm at 1st month, 318.82µm at 2nd month, 301.83µm at 3rd month and 295.17µm at 4th month (p=0.0001). (Table-III)

In the sub group analysis patients with DME the mean CMT decreased from 374.23 µm to 326.11(p=0.0001), to 295.95 (p=0.0001), to 291.88 µm (p=0.0001) at 1st month, 3rd month and 4th month respectively. In patients with RVO (CRVO and BRVO) the mean CMT decreases from 400.05 µm to 347.11(p=0.03), to 311.22(p=0.003), to 299.25(p=0.003) at 1st month, 3rd month and 4th month respectively. In patients with wet AMD the mean CMT decreases from 362.73 µm to 335.91(p=0.23). to 306.27(p=0.03), to 300.2(p=0.03) at 1st month, 3rd month and 4th month respectively. (Graph-V)

**Graph V:**
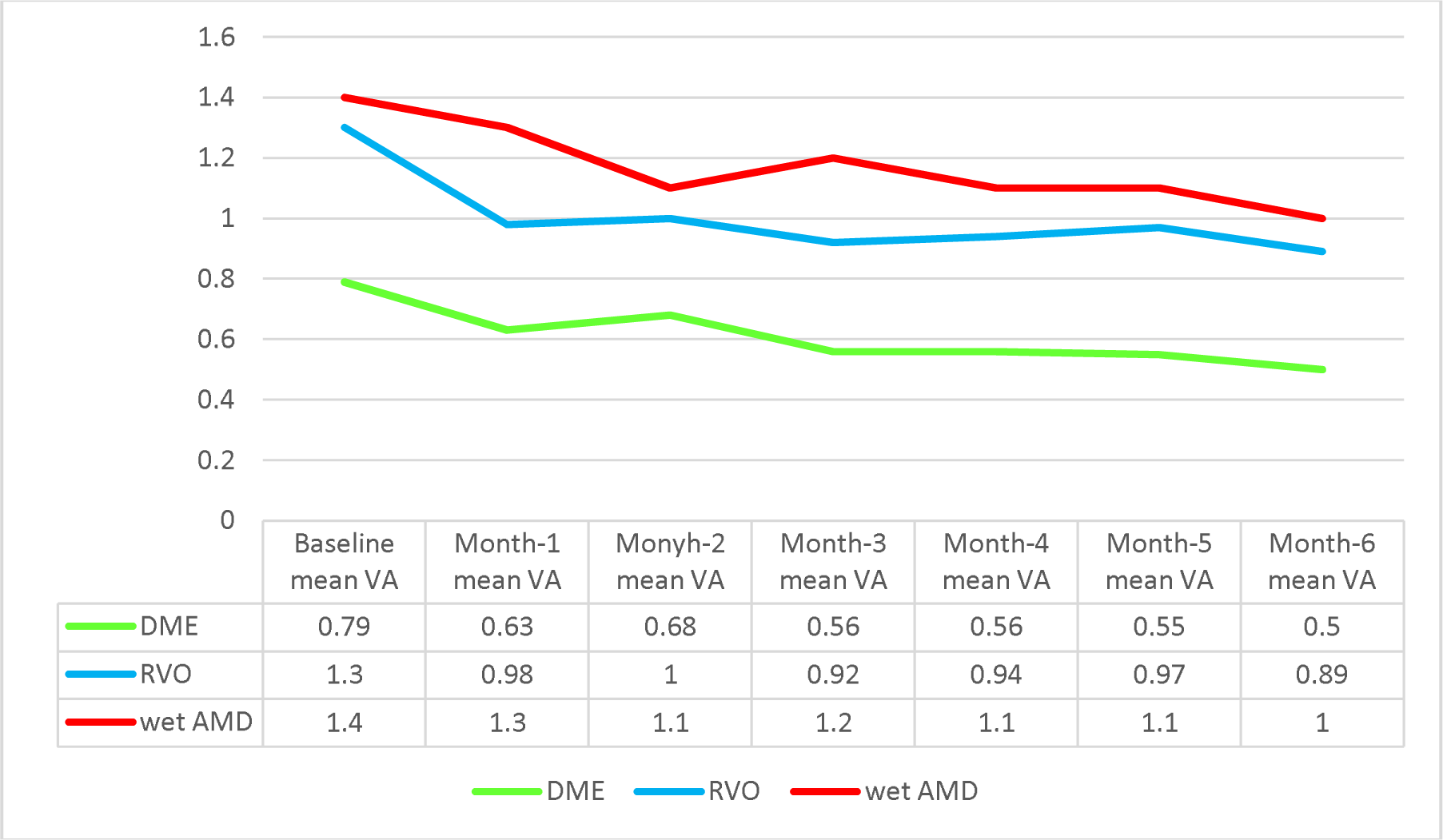
Baseline and post IVA injection mean VA of each indication in logMAR of study eyes that were candidates for IVA injection at UoG tertiary eye care and training center retina sub-specialty clinic, Gondar, Ethiopia, 2023.

**Graph VI:**
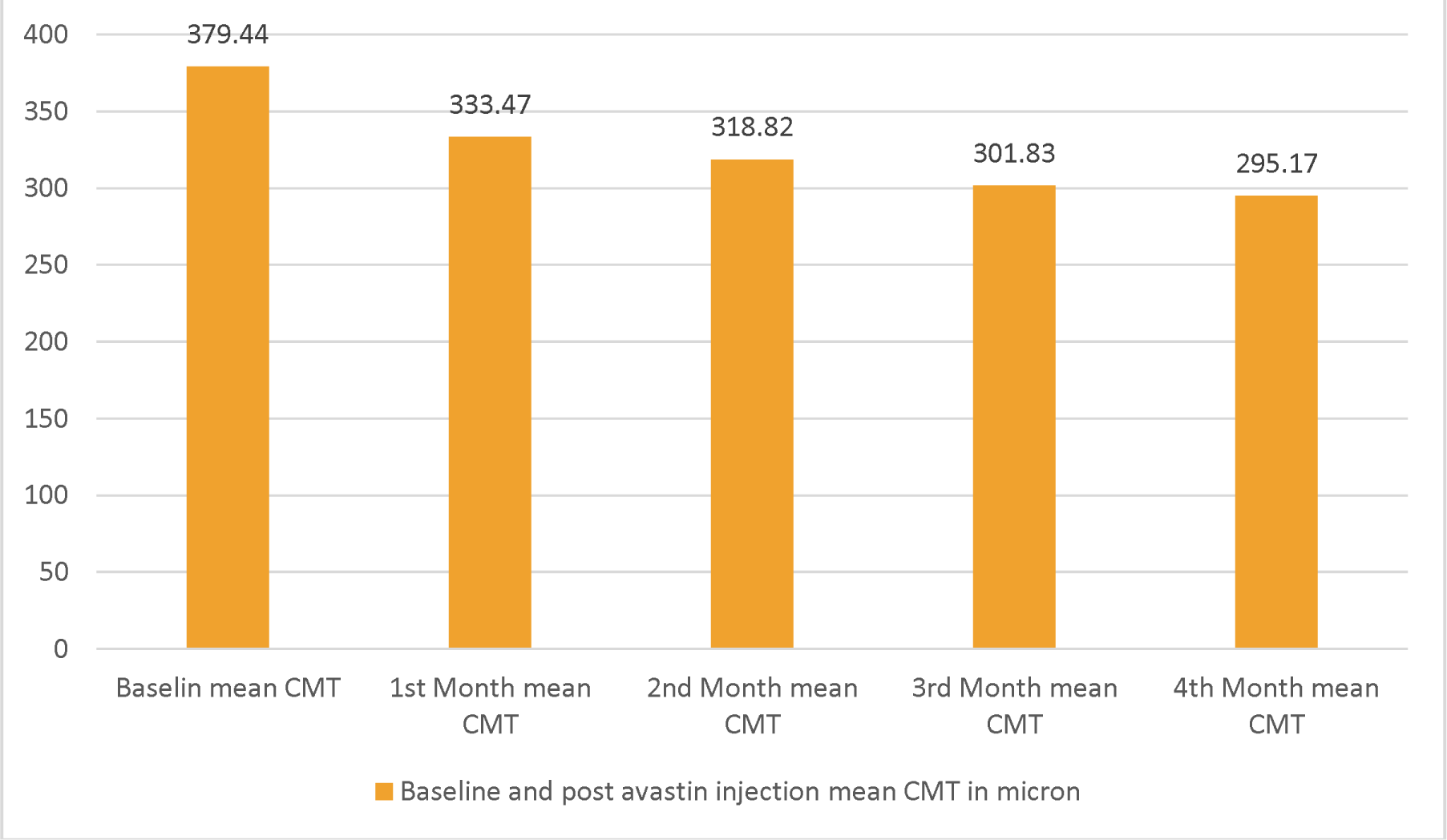
Baseline and pos IVA injection mean CMT of study eyes that were candidates for IVA injection at UoG tertiary eye care and training center retina sub-specialty clinic, Gondar, Ethiopia, 2023.

At the end of 6 month follow up 34 eyes (60%) mean VA was improved by one line and above from baseline, 16 eyes (28%) remained stable and 7 eyes (12%) have worsened by 1line and above from baseline VA. (Graph-VII)

**Graph VII:**
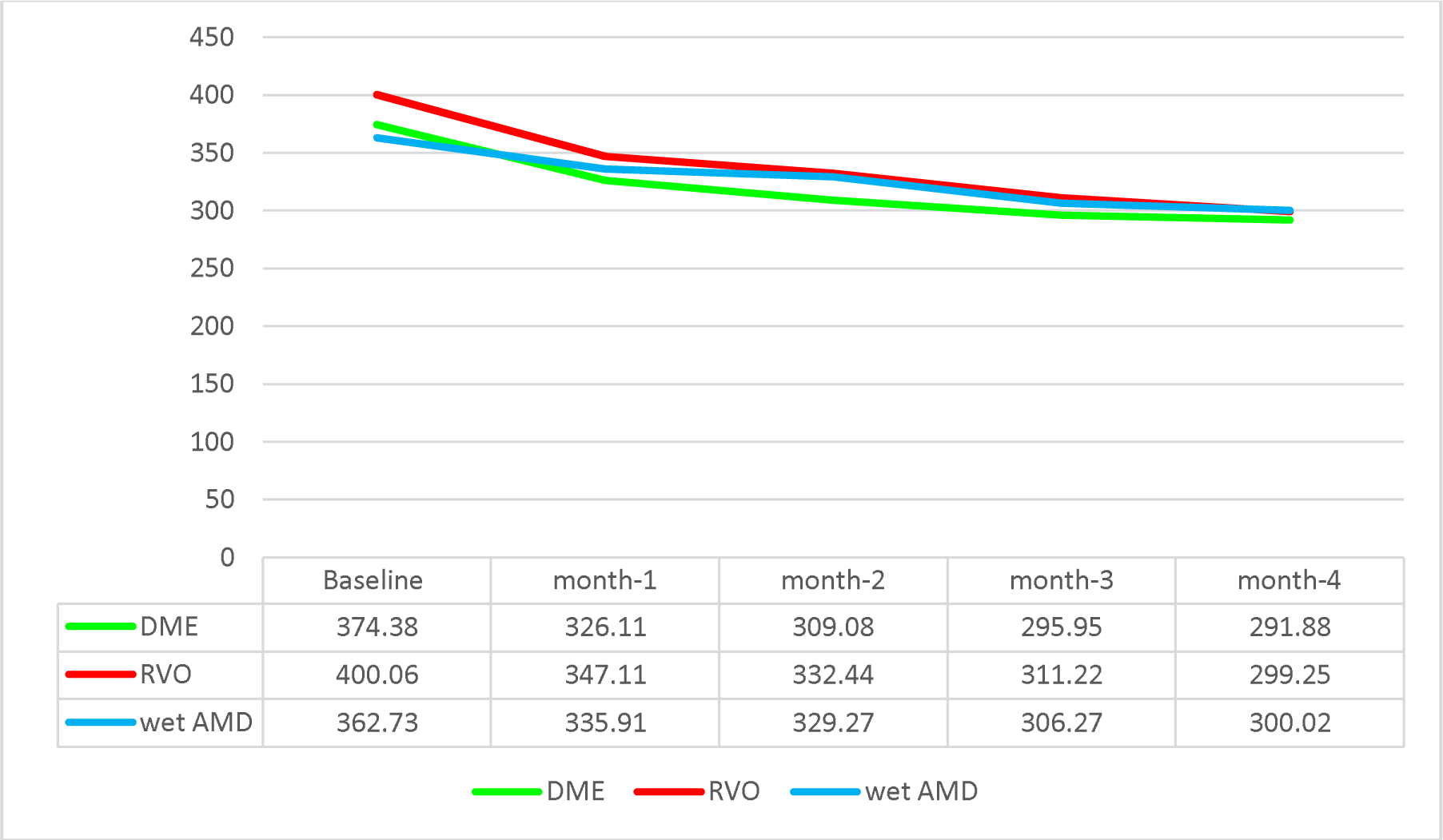
Baseline and post IVA injection mean CMT of each indication of study eyes that were candidates for IVA injection at UoG tertiary eye care and training center retina sub-specialty clinic, Gondar, Ethiopia, 2023.

In the sub group analysis and at the end of follow up (6 month), out of 31 eyes with DME 17(54.8) eyes improved from baseline, 11(35.5%) eyes remain stable and 3 (9.7%) eyes worsened from baseline. Among 16 eyes with RVO 12(75%) eyes improved from baseline, 3(18.8%) eyes remain stable and 1 (6.2%) eye worsened from baseline. In wet AMD (10 eyes); 5(50%) eyes improved from baseline, 2(20%) remain stable and 3(30%) eyes worsened from baseline. (Graph-VIII)

**Graph VIII:**
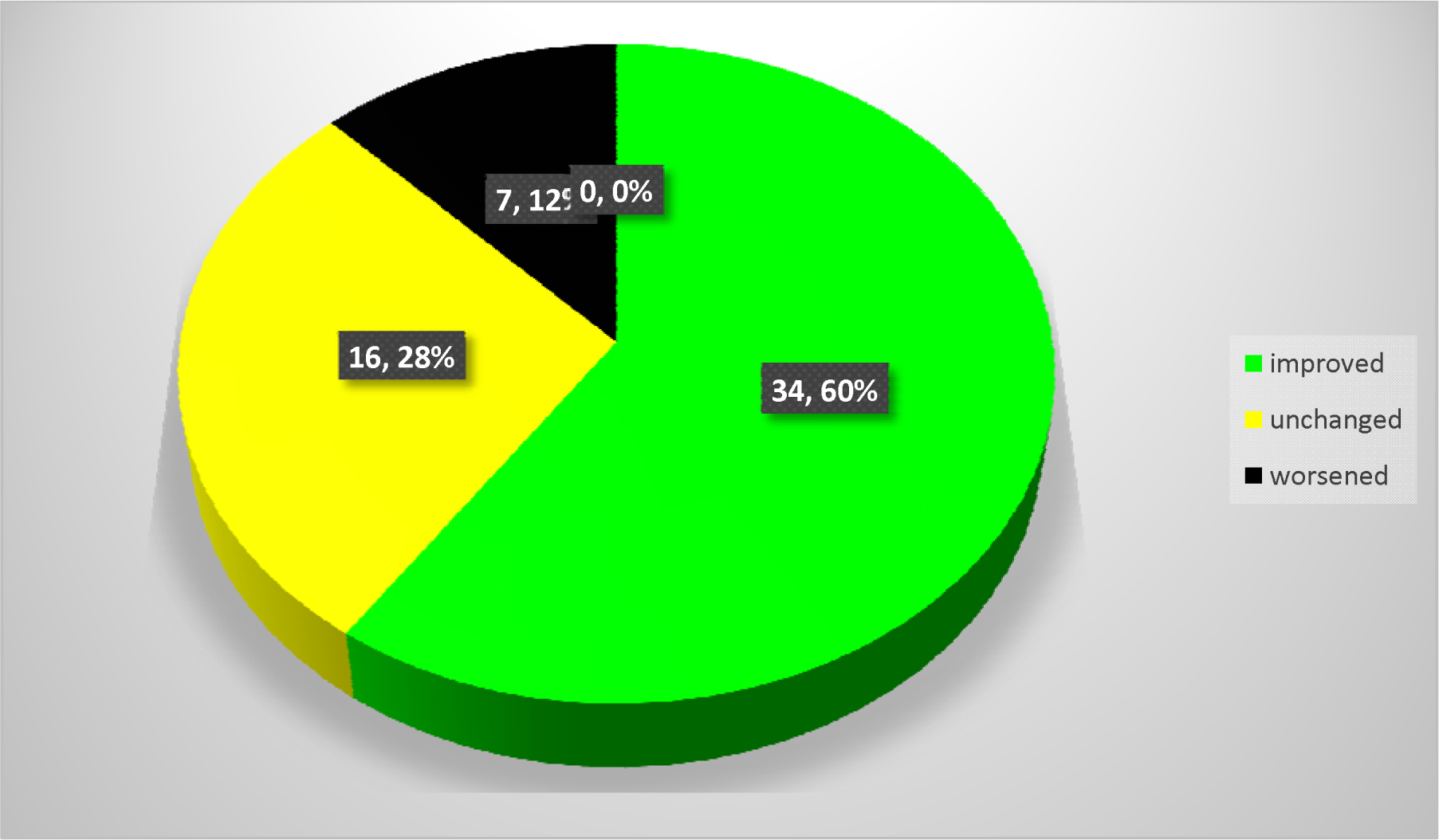
Pie chart showing the number of study eyes with their final visual outcome at UoG tertiary eye care and training center retina sub-specialty clinic, Gondar, Ethiopia, 2023.

The final mean CMT analysis at 4th month demonstrated that from 58 eyes 43 eyes (74%) improved from baseline (edema decreased by more than 10 % from baseline), 11(19%) eyes remained stable, 4 eyes (7%) had worsened CMT from baseline. (Graph-IX)

**Graph-IX:**
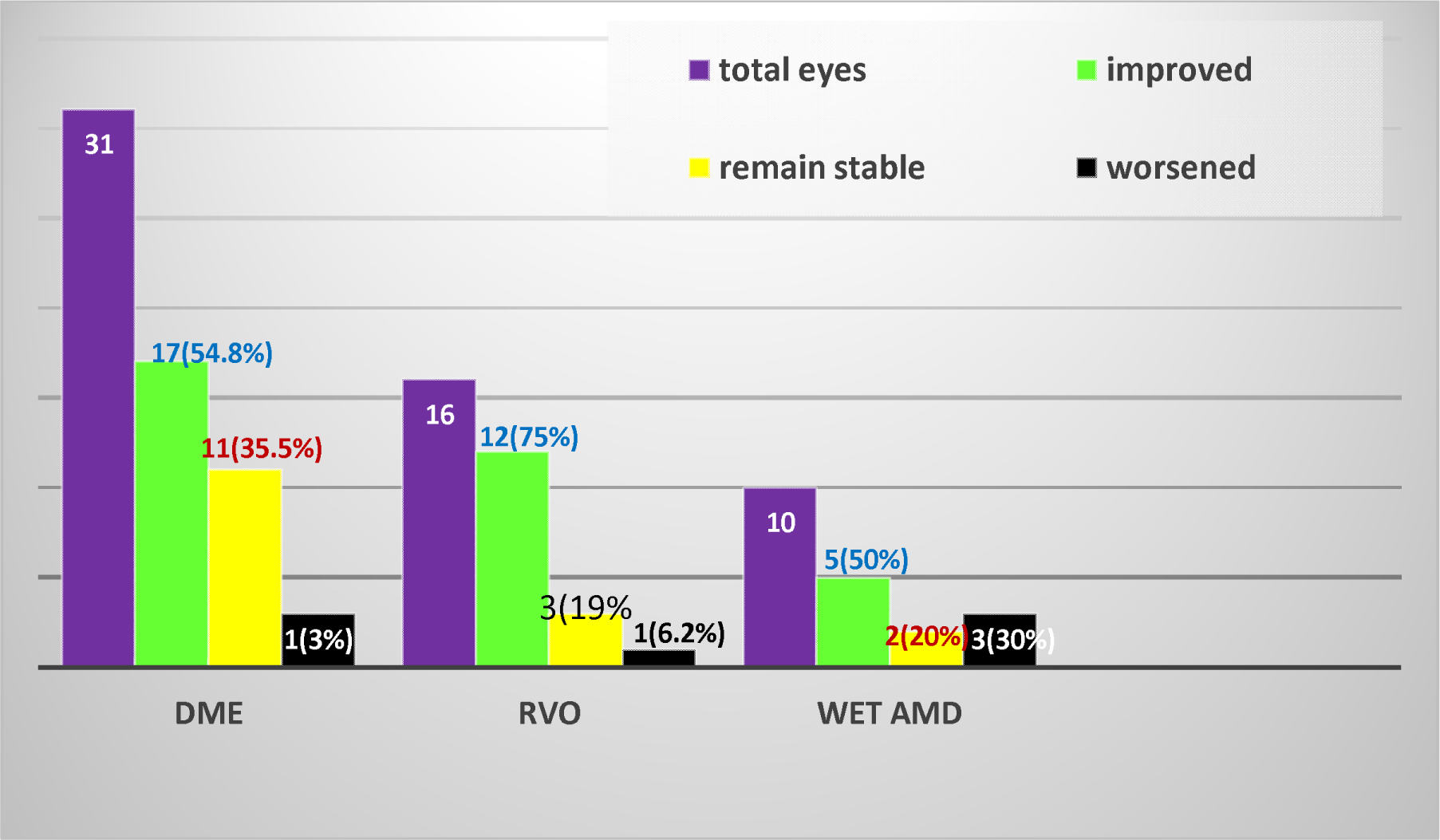
Bar graph showing the number of study eyes with final visual outcome of specific indications at UoG tertiary eye care and training center retina sub-specialty clinic, Gondar, Ethiopia, 2023.

In the sub group analysis and at the end of 4^th^ month follow up, 23(69.7%) of 33 eyes with DME, 13(81%) of 16 eyes with RVO and 6(66.7%) of 9 eyes with wet AMD improved by more than 10% from baseline mean CMT after Avastin injection and 1 eye from DME, 2 eyes from RVO and 1 eye from wet AMD had worsened CMT. (Graph-X)

**Graph X:**
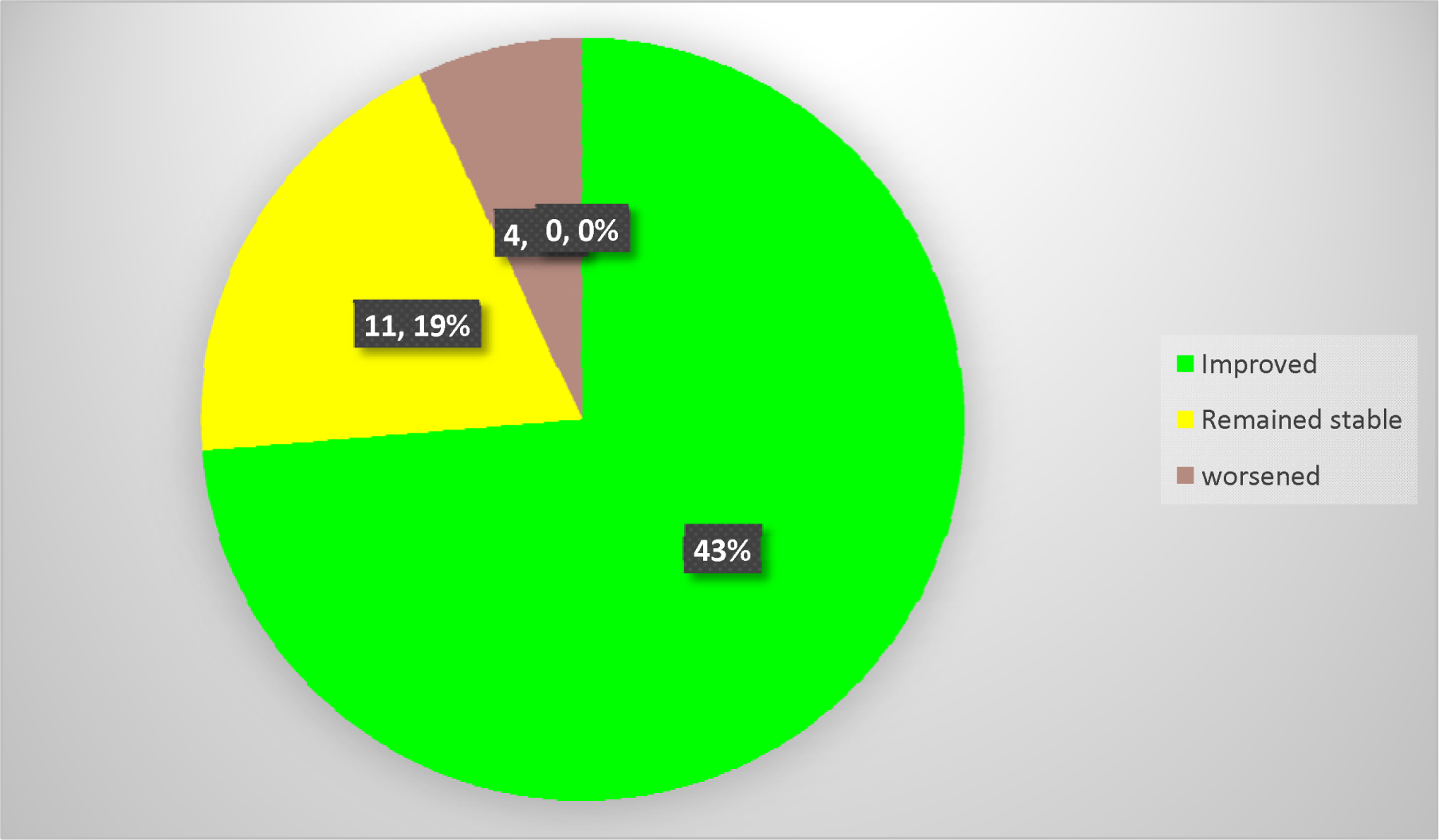
Pie chart showing the number study eyes with final anatomical outcome at UoG tertiary eye care and training center retina sub-specialty clinic, Gondar, Ethiopia, 2023.

**Graph XI:**
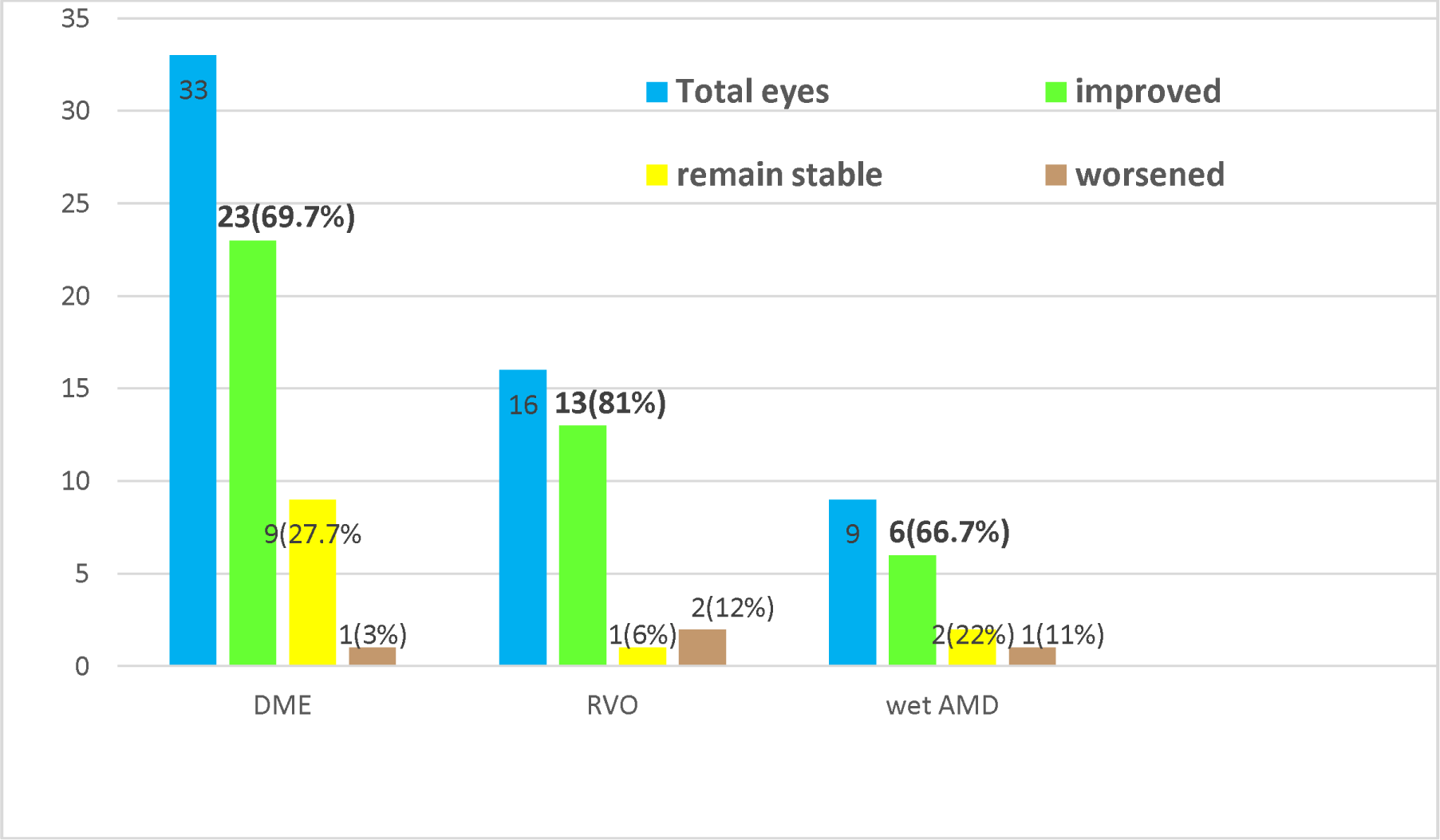
Bar graph showing the number of study eyes with final anatomical outcome of specific indications at UoG tertiary eye care and training center retina sub-specialty clinic, Gondar, Ethiopia, 2023.

### Prognostic factors for change in visual acuity

In this study only baseline mean VA had significant association with change in mean VA which means patients with better baseline VA had better VA improvement. Other factors like age, sex, mean FBS, CME, and baseline mean CMT had no significant association with change in mean VA. (Table-IV)

**Table IV:** The impact of clinical factors for change in the mean VA of injected eyes at UoG tertiary eye care and training center retina sub-specialty clinic, Gondar, Ethiopia, 2023.

| Factors | Regression Coefficient (B) | P value |
| --- | --- | --- |
| AGE | 0.122 | 0.42 |
| SEX | 0.28 | 0.66 |
| Baseline mean VA | 0.597 | 0.0001 |
| Mean FBS | -0.088 | 0.54 |
| CME | -0.099 | 0.687 |
| Diffuse ME | -0.039 | 0.87 |
| SR fluid | -0.015 | 0.92 |
| Baseline mean CMT | 0.226 | 0.086 |
| HTN | -0.141 | 0.37 |
| Number of injections | -0.008 | 0.95 |

In the sub group analysis for patients with DME and RVO baseline mean VA had significant association with change in the mean VA (p 0.017 and 0.001 respectively) other factors had no significant association.

For wet AMD there was no any factor having significant association with change in mean visual acuity.

### Prognostic factors for change in the mean CMT

Our study showed that change in mean CMT was positively affected by the number of injections (p=0.009) diffuse ME (P=0.03) and baseline CMT (p=0.007). Other factors did not have any significant association in the change in CMT. (Table-V)

**Table V:**
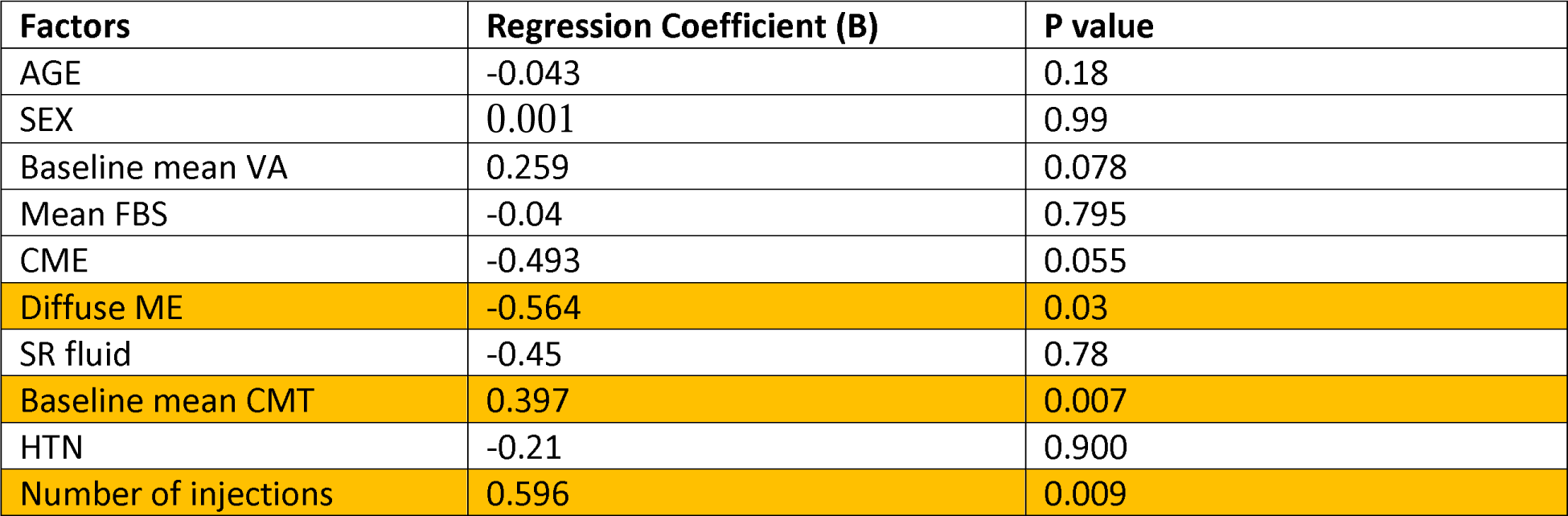
Impact of clinical factors for mean change in CMT of injected eyes at UoG tertiary eye care and training center retina sub-specialty clinic, Gondar, Ethiopia, 2023.

| Factors | Regression Coefficient (B) | P value |
| --- | --- | --- |
| AGE | -0.043 | 0.18 |
| SEX | 0.001 | 0.99 |
| Baseline mean VA | 0.259 | 0.078 |
| Mean FBS | -0.04 | 0.795 |
| CME | -0.493 | 0.055 |
| Diffuse ME | -0.564 | 0.03 |
| SR fluid | -0.45 | 0.78 |
| Baseline mean CMT | 0.397 | 0.007 |
| HTN | -0.21 | 0.900 |
| Number of injections | 0.596 | 0.009 |

In the subgroup analysis, for patients with DME mean CMT reduction had positive significant association with baseline mean CMT (P=0.0001) and the number of injections (p=0.001). Other factors had no significant association with mean CMT reduction. In patients with RVO only baseline mean CMT had significant (p=0.013) positive association with CMT reduction.

In patients with wet AMD only the number of injections had significant (p=0.03) positive association with CMT reduction. Other factors had no significant association.

## Discussion

This is the first study done to evaluate the safety and efficacy (both anatomical and functional outcome) of IVA in Ethiopia. The commonest indications of IVA injection in this study were DME followed by RVO with ME which accounted 56% and 27% of patients respectively. This is comparable to the result found in Uganda in which DME and RVO with ME accounted 53% and 25% of the indications respectively (1).

There was no any documented ocular or systemic complication after IVA injection among study patients which is similar to studies done in Uganda, Egypt, Japan and Gondar (Ethiopia) (1, 3,16,17).

Visual acuity was assessed monthly and mean VA improvement was seen starting from the first month after IVA injection and the improvement was from 1.00 to 0.8 logMAR (95%CI, 05-0.34; p=0.007) and this was maintained at second month follow up. At the third month, mean VA improved from baseline 1.00 logMAR to 0.7 logMAR (95%CI, 0.12-0.41; p=0.001). This significant change of mean VA at the 3rd month was maintained throughout the 6-months follow-up period.

In the subgroup analysis of our patients with DME the mean VA improved from baseline 0.79logMAR to 0.63 (P=0.082) at first month, 0.56 (p=0.013) at third month and 0.5 (p=0.002) at sixth month post injection.

This is comparable to the result found at Pan-American collaborative Retina Study Group done in 88 patients of 110 eyes with DME, the mean BCVA was improved from 0.8 logMAR to 0.6 logMAR(p=0.001) at the end of 6 months follow up (5).

In patients with RVO (CRVO and BRVO) the mean VA improved from baseline 1.3logMAR to 0.98(p=0.012) at first month, 0.92(p=0.008) at third month and 0.89(p=0.004) at sixth month after injection.

Similar result was found in Israel done on 35 eyes of CRVO with ME where the mean VA improved from baseline 0.9 logMAR (p=0.01) to 0.7 logMAR (p=0.01) at 3rd month and maintained for 6 month follow up(4) and in Philadelphia, USA, done on 79 eyes with wet AMD the mean BCVA improved from 0.7logMAR to 0.6logMAR at first month and maintained for 3 months (8).

In contrary to this, a study done in Germany on 21 eyes of CRVO with ME and after mean number of injection of 3.7, mean BCVA didn’t change despite a significant change in mean CMT at the end of 1 year study period (9). But this study had a follow up period of one year which may explain the observed difference with our results.

On the other hand in patients with wet AMD mean VA was improved from baseline 1.4logMAR to 1.3(p=0.7) first month, 1.2 (p=0.33) at third month and 1.0(p=0.33) at sixth months.

At the end of 6-month study period VA analysis demonstrated that from 31 eyes of DME, 17 eyes (54.8%) improved, 11 eyes (35.5%) remained stable and 3 eyes (9.7%) have worsened VA from baseline acuity.

This is comparable to a study done in Nepal on 52 eyes of DME in whom final BCVA was stable in 25 eyes (48.07), improved in 22 eyes (42.3%) and worsened 5 eyes (9.6%) ((19). In contrast to our result, in Singapore which was done on 35 eyes with DME the result showed that 13 (37.2%) eyes improved, 12 (34.3%) eyes unchanged and 10 (28.5%) worsened. But this study had smaller sample size and different methodology than ours and that may explain the observed difference.

At the end of 6-month study period out of 16 eyes of our study patients with RVO, 75% had VA improvement. This is similar to study done in Egypt at which from 26 eyes with RVO 84% of eyes had VA improvement (p=0.001) (3).

In our study there was continuous decrement of CMT from baseline in the follow up period. The mean baseline CMT was 379.44µm. After IVA injection the mean CMT decreased to 333.47µm at 1st month (95%CI,27.4-64.5; p=0.0001), 318.82µm at 2nd month (95%CI,40-81; p=0.0001), 301.83µm at 3rd month (95%CI,56-99; p=0.0001) and 295.17µm (95%CI, 77-185; p=0.0001) at 4th month.

In the sub group analysis patients with DME the mean CMT decreased from 374.23 µm to 326.11(p=0.0001), to 295.95 (p=0.0001), to 291.88 µm (p=0.0001) at 1st month, 3rd month and 4th month respectively.

This is comparable to the results found in multicenter study done in Venezuela, Mexico, Brazil, Costa Rica and USA in 110 eyes of DME, the mean baseline CMT was 387.0±182.8 µm and after injection mean CMT decreased to 287.9µm, 282.8 µm and 275.7 µm (P=0.001)at 1^st^ month, 3^rd^ moth and 6^th^ month respectively (5).

There was also continuous decrement of mean CMT in a study done in Nepal on 52 eyes of DME where the mean CMT was 449.03μm at baseline and it decreased significantly to 345.76 µm, 344.55 µm and 326.51 µm (p=0.0001) at 6 weeks, 3 months and 6 months post injection, respectively (13).

Further subgroup analysis of our patients with RVO showed the mean CMT decreased from 400.05 µm to 347.11(p=0.03), to 311.22(p=0.003), to 299.25(p=0.003) at 1st month, 3rd month and 4th month respectively.

This was also similar result found in a study done in India on 21eyes with RVO with ME, the mean baseline CMT was 647.81µm and decreased to mean 393.43µm at one month and 320.90µm (p=0.0001) at 3rd month and another study in Germany done on 21 eyes of CRVO with ME, where the mean CMT was 780µm baseline, 507µm at 6 months, and 462µm at 12 months (12,20).

In another subgroup analysis of our patients with wet AMD the mean CMT decreased from baseline 362.73 µm to 335.91(p=0.23) at first month 306.27(p=0.03), third month and 300.2(p=0.03) at fourth month post injection.

Similar result was also found in Romania done on 117 eyes with wet AMD where baseline mean CMT was 450.94 µm and three months after injection mean CMT decreased to 309.13 µm (11).

The mean CMT analysis at 4^th^ month demonstrates that from 33 eyes with DME 23 eyes (69.7%) had significant improvement from baseline mean CMT (P=0.0001). In contrast to our study, the multicenter study in Venezuela, Mexico, Brazil, Costa Rica and USA in 110 eyes of DME, 55% of study eyes had improvement in CMT at the end of the study period (21). This may be due to the differences in the sample size.

At the end of 4th month from 16 eyes with RVO, 81% of eyes had mean CMT reduction (p=0.003). This is similar to a study done in USA on 30 eyes with RVO, 86.7% of eyes had mean CMT reduction (p=0.001) (14).

In our study, only the mean baseline VA had significant association with the change in mean VA, which means the better the mean base line VA, the better the final visual improvement. Other factors like age, sex, mean FBS, type of macular edema and baseline mean CMT had no significant association with change in mean VA.

In the sub group analysis for patients with DME and RVO baseline mean VA had significant association with change in mean VA with p=0.017) and p=0.00 respectively.

This is similar to studies done in Brazil, India, Korea and Poland which found that age, sex, FBS and blood pressure did not have association with change in BCVA following IVA injection (22,23,2). Similar studies done in India, UK, Singapore and Germany also showed that the base-line mean CMT had no effect on visual outcome (25,26,27,28,)

Two studies done in Taiwan and France on patients with RVO reported that the higher the baseline VA the better the VA response after injection (29,28).

In contrast to our results, studies done in India, Thailand, USA on patient with DME and a study done in Korea on wet AMD patients found that the lower the baseline VA the better the visual out (22,23,31,32).

Additional contradictory results from ours are reports from Germany, France, and Korea on patients with RVO and a study done in Thailand on DME patients which found that age and type of macular edema (sub-retinal fluid and cystoid macular edema) were strong prognostic factors for visual outcome. The younger the age the better the visual outcome, the thicker SRF and presence of CME had worst VA outcome after IVA injection (27,29,31,33).

This study showed that change in CMT had strong positive association with the number of injections (p=0.009) and baseline CMT (p=0.007). This means patients with more number of injections and thicker baseline CMT had significant post injection improvement in CMT. Other factors had no significant association in the change in CMT. Further subgroup analysis of patients with DME, RVO and AMD also showed similar association.

Studies done in USA, Palestine and Uganda also found that higher baseline mean CMT was significantly associated with reduction mean CMT after IVA injection (1, 26, 32).

However, a study done in Germany on 32eyes of BRVO with macular edema reported that baseline mean CMT was not associated with post injection CMT reduction (23). Longer study period may explain this difference.

## Conclusion

This study showed that IVA injection had significant effect on improving mean visual acuity and mean CMT reduction. Improvement in vision was more marked among patients with DME when compared to other causes of ME. Thicker baseline central macula and the number of injections were strong predictor of CMT reduction whereas a better baseline vision was a strong predictor of visual improvement at the end of follow up. IVA injection had no any identified ocular and systemic complications.

Although this is the first study to evaluate the safety and efficacy (both functional and anatomical) of IVA in Ethiopia, the relatively small sample size and the retrospective study design may have limited the results.

## Data Availability

All data produced in the present study are available upon reasonable request to the authors

